# Macrophage Therapy for Acute Liver Injury (MAIL): a Phase 1 Randomised, Open-Label, Dose-Escalation Study to Evaluate Safety, Tolerability, and Activity of Allogeneic Alternatively Activated Macrophages in Patients with Paracetamol-induced Acute Liver Injury

**DOI:** 10.1101/2024.05.28.24306936

**Authors:** Christopher Humphries, Melisande Addison, Guruprasad Aithal, Julia Boyd, Lesley Briody, John DM Campbell, Maria Elena Candela, Ellise Clarke, James Coulson, Nicholas Downing-James, Robert John Fontana, Ailsa Geddes, Julia Grahamslaw, Alison Grant, Anna Heye, James A Hutchinson, Ashley Jones, Fiona Mitchell, Joanna Moore, Alice Riddell, Aryelly Rodriguez, Angela Thomas, Garry Tucker, Kim Walker, Christopher J Weir, Rachel Woods, Sharon Zahra, Stuart J Forbes, James Dear

**Affiliations:** Centre for Cardiovascular Science, University of Edinburgh, The Queen’s Medical Research Institute, 47 Little France Crescent, Edinburgh, EH16 4TJ, UK; Centre for Regenerative Medicine, University of Edinburgh, 5 Little France Drive, Edinburgh, EH16 4UU, UK; MAIL Trial Data Monitoring Committee, Edinburgh Clinical Trials Unit, Usher Institute, University of Edinburgh, 3 Little France Road, Edinburgh, EH16 4UX, UK; NIHR Nottingham Biomedical Research Centre, Nottingham University Hospitals NHS Trust, Queen’s Medical Centre, Derby Road, Nottingham, NG7 2UH, UK; Edinburgh Clinical Trials Unit, Usher Institute, University of Edinburgh, 3 Little France Road, Edinburgh, EH16 4UX, UK; Edinburgh Clinical Research Facility, Royal Infirmary of Edinburgh, 51 Little France Crescent, Edinburgh, EH16 4SA, UK; Scottish National Blood Transfusion Service, Jack Copland Centre, 52 Research Avenue North, Edinburgh, EH14 4BE, UK; Emergency Medicine Research Group Edinburgh, Royal Infirmary of Edinburgh, 51 Little France Crescent, Edinburgh, EH16 4SA, UK; MAIL Trial Steering Committee, Edinburgh Clinical Trials Unit, Usher Institute, University of Edinburgh, 3 Little France Road, Edinburgh, EH16 4UX, UK; Centre for Precision Cell Therapy for the Liver, Lothian Health Board, Queens Medical Research Institute, 47 Little France Crescent, Edinburgh, EH16 4TJ UK

## Abstract

**Introduction:** Acute Liver Failure (ALF) has no effective treatment other than liver transplantation, and is commonly caused by paracetamol overdose. New treatments are needed to treat and prevent ALF. Alternatively activated macrophages (AAMs) can promote resolution of liver necrosis and stimulate hepatocyte proliferation. Using AAMs in unscheduled care requires the use of an allogeneic product. A clinical trial is needed to determine the safety and tolerability of allogeneic AAMs.

**Methods and analysis:** A single centre, open-label, dose-escalation, phase 1 randomised trial to determine whether there is dose-limiting toxicity of AAMs in patients with paracetamol-induced acute liver injury. Randomisation will occur at higher doses.

**Ethics and dissemination:** The trial will be conducted according to the ethical principles of the Declaration of Helsinki 2013 and has been approved by North East – York Research Ethics Committee (reference 23/NE/0019), NHS Lothian Research and Development department, and the UK Medicines and Healthcare products Regulatory Agency. When the trial concludes, results will be shared by presentation and publication.

Trial registration number: ISRCTN 12637839.

## Introduction

### Background

Acute liver failure (ALF) has no effective treatment other than liver transplantation, which has associated morbidity/mortality, expense and a scarcity of donor livers.(1) The commonest cause of ALF is paracetamol (acetaminophen) overdose (POD). In the UK, around 50,000 people need emergency treatment with acetylcysteine (n-acetylcysteine – NAC), every year.(2) The USA has around 89,000 hospital visits annually for POD, with an estimated annual treatment cost of US$1billion.(3) Around 10% of these patients develop biochemical evidence of acute liver injury (ALI) and around 1 in 200 develop ALI severe enough after POD to need a liver transplant.(4) Mortality in the ALF group is around 30-35%, with about 30% receiving a liver transplant (most of whom survive but will require lifelong ongoing treatment and often have morbidity secondary to the immunosuppression required for transplantation).(1) Many of these patients occupy critical care beds for weeks. Aside from paracetamol, there is a wide range of other causes for ALF including non-paracetamol drug-induced liver injury and viral infection. There is no specific treatment for ALF due to non-paracetamol causes (NAC is sometimes used, but has modest efficacy).(5)

In cases of POD, the metabolic pathways that eliminate the drug become saturated and a highly reactive metabolite, N-acetyl-p-benzoquinone imine (NAPQI), is made in excess by the cytochrome P450 enzyme system. NAPQI is directly toxic to hepatocytes and leads to liver necrosis.(6) The immune system responds by a massive infiltration of circulating inflammatory monocytes into the liver.(7) These cells create a highly toxic inflammatory environment. In humans, the degree of infiltration of monocytes, identified by concurrent monocytopenia in the circulation, reflects the degree of ALI.(8) Over a period of 24-48 hours, liver-infiltrating monocytes differentiate to macrophages which adopt an anti-inflammatory, wound-healing phenotype (“M2”)(7). These macrophages initiate the healing process and facilitate the transition from the initial inflammatory phase of ALF to the resolution phase. Macrophage-mediated resolution of ALI comprises phagocytosis of necrotic hepatocytes, induction of regeneration by proliferation of hepatocyte progenitors and suppression of systemic inflammation via an anti-inflammatory cytokine effects.(9)

Currently, the only effective treatment for preventing ALI after POD is NAC. If treatment is commenced within 8 hours of overdose, then NAC is near 100% effective. However, effectiveness drops substantially when treatment is delayed; NAC is near ineffective when treatment is delayed greater than 20 hours after overdose, though delayed treatment may reduce the subsequent development of hepatic encephalopathy and mortality. (10,11) There is a clear unmet need for new treatments to prevent and/or treat ALF.

### Rationale for study

We urgently need a treatment which prevents the progression of ALI to ALF to prevent liver transplantation or death. Halting or reversing progression to ALF would be life-saving and remove the need for immunosuppression and morbidity/mortality associated with transplantation. The proposed solution is to develop allogeneic alternatively-activated macrophages (AAMs) as a treatment. An allogeneic product is necessary for the acute setting, when autologous monocytes may have altered properties, and there is insufficient time to produce autologous cell products. This cell therapy has clear efficacy when delivered after liver injury is established in mouse models.(12) This is because macrophage therapy reverses injury rather than preventing it. Early phase human studies of chronic liver disease using autologous macrophage therapy have provided evidence to support the safety of AAMs, but a phase 1 trial is required to establish whether an allogeneic product is feasible.(13)

The primary aim of this trial is to determine the safety and tolerability of AAMs. There are multiple causes of ALF other than paracetamol, but POD is the most common cause in the United Kingdom and patients typically have a clearly defined onset of injury and blood tests to confirm the diagnosis. Due to the distribution and mechanisms of action of AAMs, healthy volunteers are not a suitable test population. AAMs localise to injured liver tissue, where they target necrotic hepatocytes and reduce inflammatory factors. Therefore, humans with healthy livers would not provide meaningful safety or activity data.

If successful, this study will pave the way for a randomised controlled trial of the effectiveness of macrophage cell therapy in patients with paracetamol-induced ALI and facilitate the development of macrophage cell therapy for other causes of ALF. Here, we summarise the MAIL (Macrophage therapy for Acute Injury to the Liver) trial protocol in accordance with the SPIRIT guidance for randomised trial protocols and the SPIRIT-DEFINE extension for dose-finding studies.(14)

### Objectives

The primary objective of this phase 1 trial is to determine whether there is dose-limiting toxicity (DLT) produced by allogeneic AAMs in patients with paracetamol-induced ALI.

The secondary objectives are to explore the safety, activity and immunogenicity of allogeneic AAMs in patients with paracetamol-induced ALI.

### Trial design

MAIL is a single centre, open-label, dose-escalation, phase 1 randomised trial to determine whether there is DLT of AAMs in patients with paracetamol-induced ALI. Randomisation will occur at higher doses. Sentinel dosing of patients, and dose escalation decisions will be guided by an independent Data Monitoring Committee (DMC).

Figure 1a describes the planned trial design. The heterogeneity in patient outcomes following the development of paracetamol-induced ALI leads to a risk of allocation bias if the next dose of cells to be given is known by the recruiting team, as less unwell patients could be preferentially enrolled at higher doses. Therefore, we will use randomisation with concealed allocation at higher dosing levels, and will blind the recruiting team to prevent prediction of the next dose of AAM treatment.

**Figure 1a.**
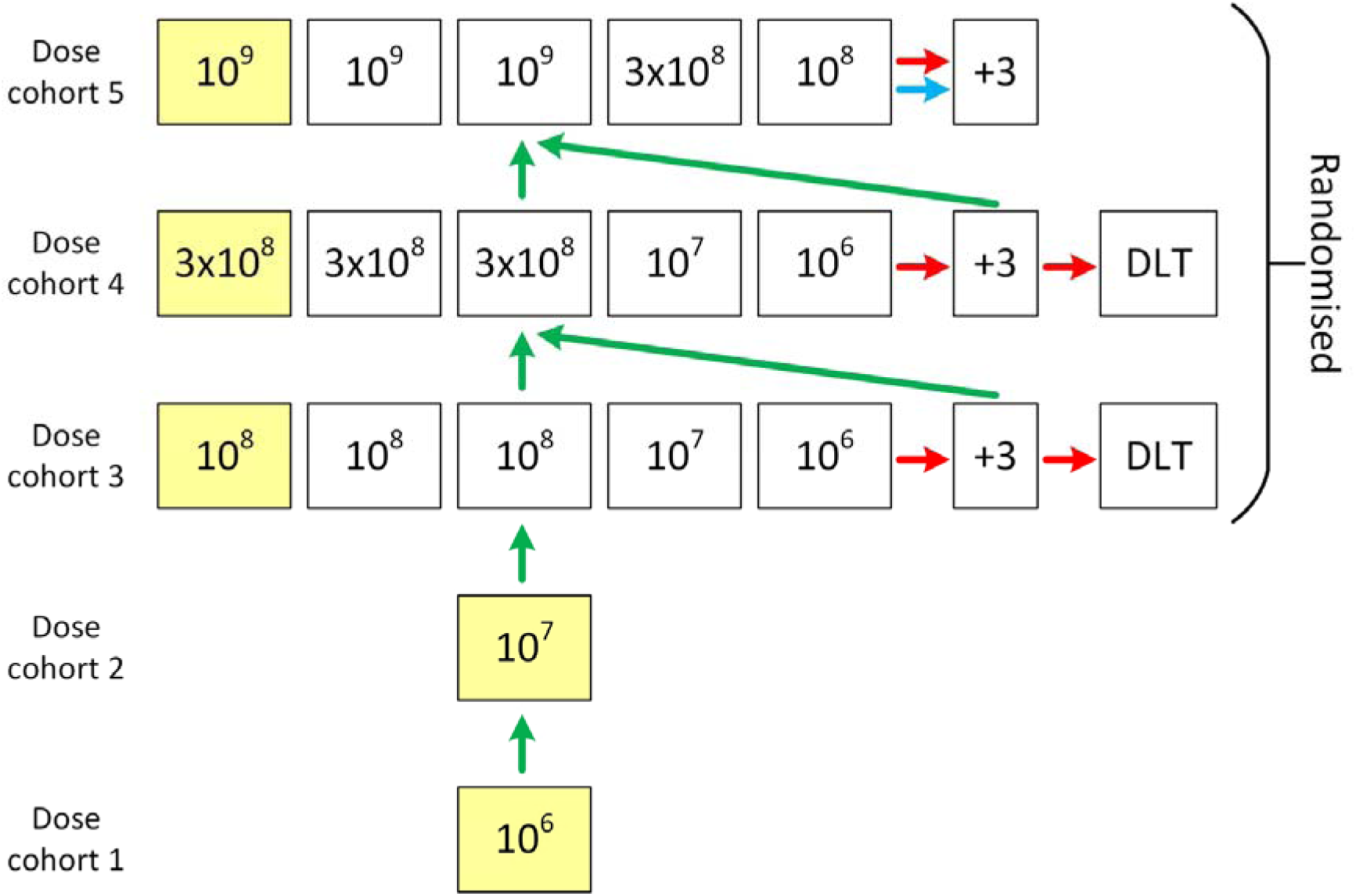
Overview of MAIL Trial design – Scenario 1 (rapid escalation). In cohort 1, the first patient will be dosed with 10 cells. In scenario 1 there is no safety concern following DMC review and the dose is escalated to 10 (cohort 2) and then into randomised blocks in cohorts 3, 4 and 5. A green arrow represents a DMC recommendation to dose escalate. A red arrow indicates a DMC recommendation to expand at that dose. The blue arrow indicates a trial team option to expand at highest dose. DLT is dose-limiting toxicity. Yellow squares are sentinel dosed patients. The highest dose is up to 10 cells.

Extrapolating from mouse models we predict the clinically active dose will be between 10^8^-10^9^ cells. The biologically active dose in mouse models of paracetamol liver injury is 10^6^ cells (the lowest trial dose). Scaling up to humans, the predicted human equivalent dose is between 2 x 10^8^ (if calculated using body surface area) to 2 x 10^9^ (using body weight). If allometric scaling is used with liver weight and blood flow as variables the human equivalent dose of AAMs is around 10^9^ (the highest trial dose).

Within each dosing cohort, the first participant to receive the highest dose will be a sentinel dosing patient. There will be at least one week pause to recruitment following the conclusion of each dosing cohort. The DMC will make a decision on dose progression using data which will include all unblinded demographic, concomitant medications, infusion, physiological, safety and activity data gathered at days -1, 0, 1, 2 (if applicable) and 7 at a minimum, which will be quality-control checked before DMC review. Data from day 30 will also be included if available at the time of the DMC meeting. The Sponsor, Chief Investigator and Trial Steering Committee (TSC) will review the DMC recommendation and make a documented decision regarding dose progression.

For all dosing cohorts the DMC can recommend a further three patients be dosed at the highest dose in that cohort. If one participant in a dose cohort experiences potential DLT, then the DMC can recommend treating three additional participants at the same dose. If DLT occurs in two or more patients in any size of dose cohort, then recruitment should end.

If there is a safety signal in dose cohorts 1 or 2, the DMC can recommend the addition of two extra patients to that cohort. If a further two patients are recommended by the DMC in cohort 2 (10^7^) but not in cohort 1 (10^6^) and no further signals are observed at 10^7^ (Scenario 2 - Figure 1b), then the 10^7^ doses are dropped from the randomisation in cohorts 3 and 4 (four patients randomised: three at higher dose and one at 10^6^).

**Figure 1b.**
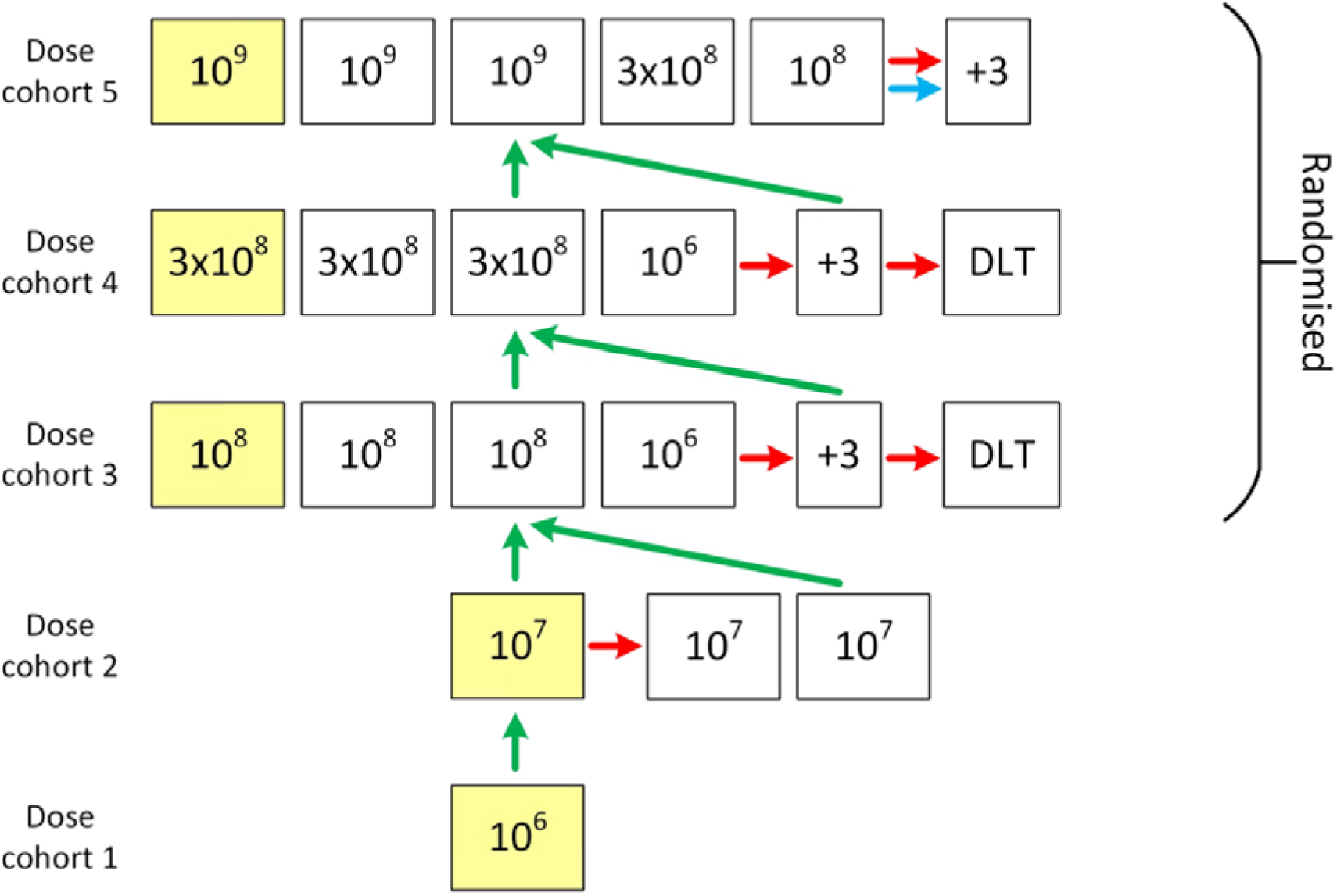
Overview of MAIL Trial design – Scenario 2. In scenario 2 there is a safety signal in cohort 2 and the DMC advise two further patients are added. No further signals are observed at 10 and the 10 doses are dropped from the randomization in cohorts 3 and, 4. A green arrow represents a DMC recommendation to dose escalate. A red arrow indicates a DMC recommendation to expand at that dose. The blue arrow indicates a trial team option to expand at highest dose. DLT is dose-limiting toxicity. Yellow squares are sentinel dosed patients. The highest dose is up to 10 cells.

If an additional two patients are recommended by the DMC in cohort 1 (10^6^), then two patients must also be added to cohort 2 (10^7^). The trial design would thereafter effectively follow a ‘3+3’ phase 1 trial design (Scenario 3 - Figure 1c).(15)

**Figure 1c.**
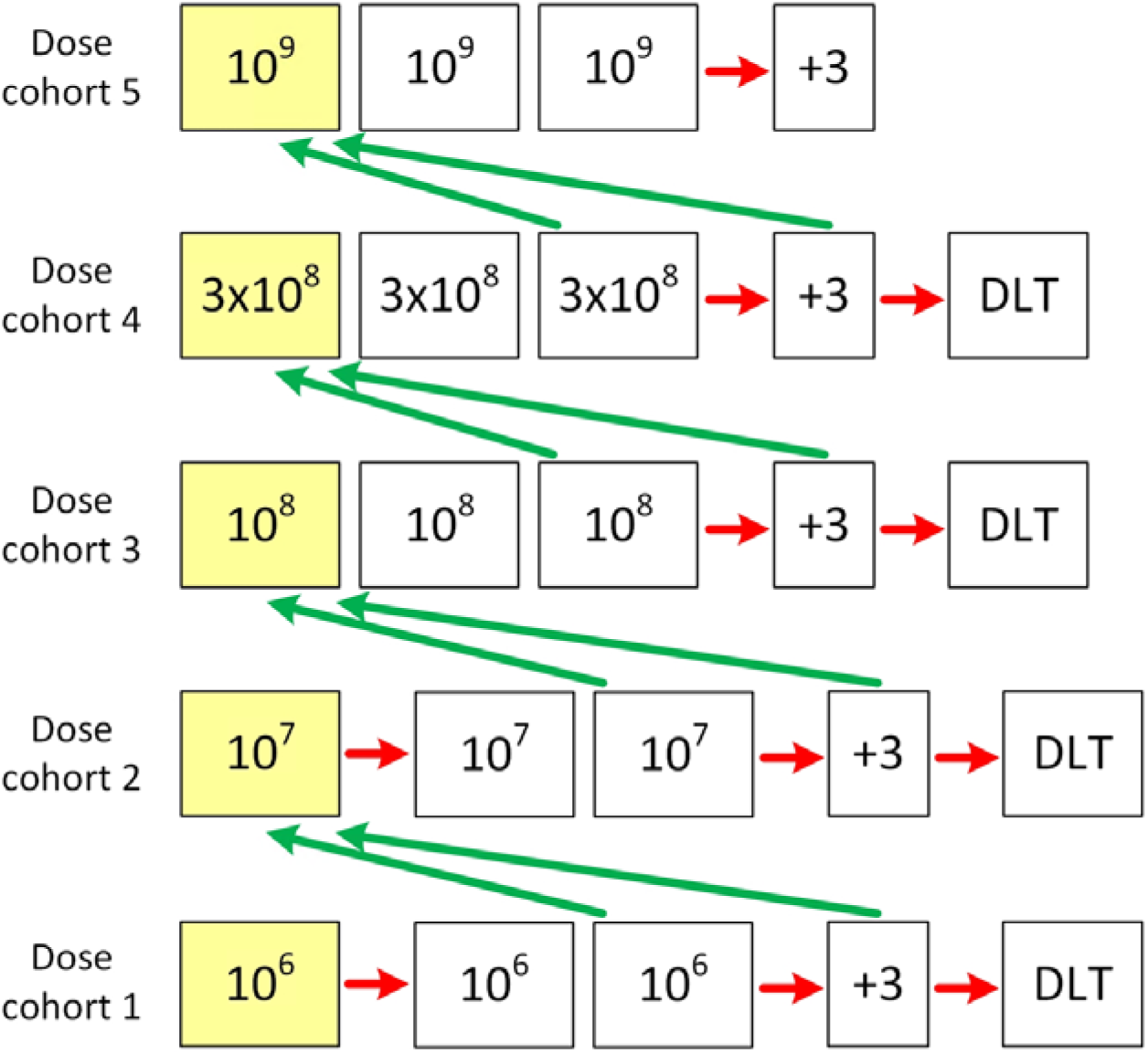
Overview of MAIL Trial design – Scenario 3 (3+3 escalation). In Scenario 3 there is a safety concern and there is expansion of cohorts 1 and 2 into 3+3 blocks. Each square represents one patient dosed at the dose indicated, except the +3 box which indicates an option to dose three further patients at the highest dose in that cohort. A green arrow represents a DMC recommendation to dose escalate. A red arrow indicates a DMC recommendation to expand at that dose. DLT is dose-limiting toxicity. Yellow squares are sentinel dosed patients. The highest dose is up to 10 cells.

## Methods

### Study oversight

The MAIL trial is an investigator-led study, funded by the Medical Research Council (reference MR/T044802/1) and sponsored by ACCORD (Academic and Clinical Central Office for Research and Development for NHS Lothian/University of Edinburgh, reference AC22087). Trial oversight is directly provided by and independent TSC and DMC, who will provide advice and review of safety. The trial has been authorised by the UK Medicines and Healthcare products Regulatory Agency (MHRA) (CTA 01384/0270/001-0001).

The composition of the TSC, Steering Group, Regulatory and Clinical Trial Group, and DMC are provided as Supplement 2.

### Study setting

This is a single-centre trial, taking place at the Royal Infirmary of Edinburgh and anticipated to run for approximately two years. Eligible inpatients who have signed the informed consent will receive a single infusion of AAMs in the Royal Infirmary of Edinburgh Clinical Research Facility (RIECRF). Patients will remain in the RIECRF for at least 6 hours after dosing, then will return to the toxicology ward or transplant unit for continued monitoring. Patients will also receive acetylcysteine as per standard care. The period of follow up will be 30 days. Please refer to Figure 2 for a study visits flow diagram.

**Figure 2.**
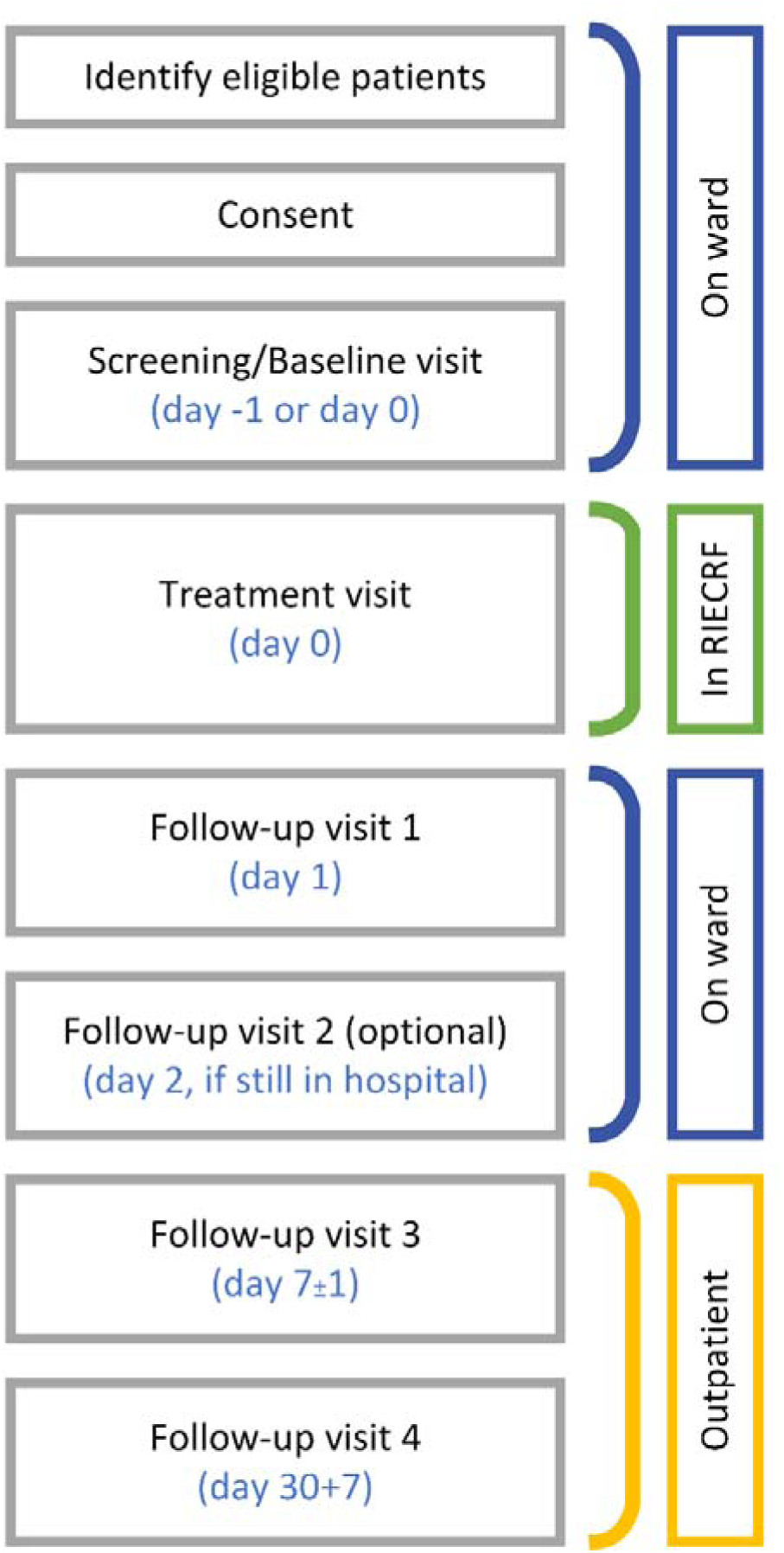
Study visits flow diagram.

### Patient and public involvement

Draft versions of the participant information sheet and lay summary were reviewed by the Edinburgh Clinical Research Facility Patient Advisory Group, a well-established Patient and Public Involvement group. They will also be asked to review lay summaries of results prior to dissemination. In addition, a member of the Sheffield Emergency Care Forum has joined the TSC as an independent lay member.

## Eligibility criteria

### Inclusion criteria

All of:

1. Serum ALT activity >1000U/L at any point between admission for the current paracetamol overdose and confirmation of eligibility.
2. History of paracetamol overdose within 5 days of ALT >1000U/L. Overdoses of paracetamol alone and mixed overdoses are eligible.
3. Other causes of ALT increase excluded based on previous investigations and trial screening. This will be documented in the patient’s medical notes.
4. Provision of written informed consent.
5. Adult male or female (16 years old or above).
6. Deemed safe for hospital discharge from a mental health perspective after full mental health assessment by a mental health professional. This will be documented in the patient’s medical notes.
7. Patients with childbearing potential must have a negative urine or serum pregnancy test at screening. If the patient is of childbearing potential or is a male with a female partner with childbearing potential, the patient, and their partner(s), must agree to use a highly effective method of contraception throughout the trial period and for 90 days post study completion.

### Exclusion criteria

Any of:

1. Patients who do not have the capacity to consent.
2. Any situation that in the Investigator’s opinion may interfere with optimal or safe study participation such as clinically relevant concurrent illness, alcohol or drug abuse, potential non-compliance, or inability to co-operate.
3. Patients with known current viral Hepatitis A (HAV), B (HBV), C (HCV) or E (HEV), Human Immunodeficiency Virus (HIV), Cytomegalovirus (CMV), Epstein-Barr Virus (EBV) or known COVID-19 infection.
4. Patients who are pregnant, or are planning on becoming pregnant during the study, or are breastfeeding and wish to continue breastfeeding. Patients of childbearing potential, or male patients with a female partner of childbearing potential, who do not agree to use a highly effective method of contraception as detailed above.
5. Patients who have previously participated in this study or another ATMP.
6. Potentially life-threatening liver injury with an immediate need for transplantation as documented in the patient’s medical notes.
7. Patients that are listed for or previously received any organ transplant.
8. Patients with stage 4 or 5 chronic kidney disease.
9. Any history of or suspected hypersensitivity to the cell product, excipients, or possible residual components used in manufacture.
10. Patients who are currently enrolled in another ATMP or CTIMP.

## Interventions

The product under investigation is an allogeneic, AAM cell therapy given at doses from 10^6^ up to 10^9^. The apheresis product will be manufactured by the Scottish National Blood Transfusion Service using healthy volunteer donations at a licensed site using methodology described previously.(16) The product has received manufacturing approval from the MHRA. The duration and rates of AAM infusion are given in table 1.

**Table 1:**
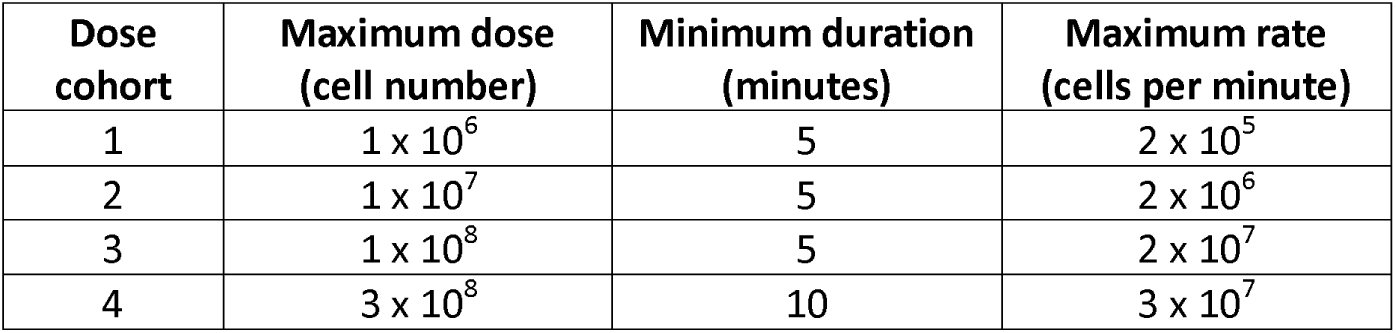
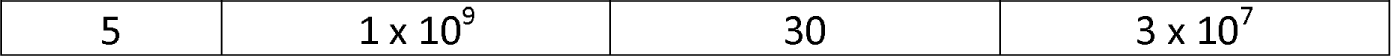
Duration and rate of intravenous AAM infusion by dose cohort.

| Dose cohort | Maximum dose (cell number) | Minimum duration (minutes) | Maximum rate (cells per minute) |
| --- | --- | --- | --- |
| 1 | $1 \times 10^6$ | 5 | $2 \times 10^5$ |
| 2 | $1 \times 10^7$ | 5 | $2 \times 10^6$ |
| 3 | $1 \times 10^8$ | 5 | $2 \times 10^7$ |
| 4 | $3 \times 10^8$ | 10 | $3 \times 10^7$ |
| 5 | $1 \times 10^9$ | 30 | $3 \times 10^7$ |

In addition to AAMs, all patients will be treated with acetylcysteine per standard clinical care. The SNAP regimen for delivering acetylcysteine will be used as recommended by the National Poisons Information Service via TOXBASE® (www.toxbase.org) and endorsed as the default standard of care by the Royal College of Emergency Medicine.(17) The SNAP regimen has a substantially lower rate of adverse drug reactions, which will facilitate the identification of treatment emergent adverse events due to the cell therapy.

Chlorphenamine 10mg will be administered intravenously approximately 30 minutes ahead of macrophage infusion to minimise the likelihood of allergic reaction. If a patient has known allergies or contraindications to chlorphenamine, 10mg cetirizine hydrochloride will be taken orally instead.

## Outcomes

### Primary outcome

DLT occurring within 30 days of infusion. DLT is defined as a clinically significant adverse event or abnormal laboratory value assessed as unrelated to disease progression, intercurrent illness, or concomitant medications and either meeting the National Cancer Institute (NCI) Common Terminology Criteria for Adverse Events Grade 3, 4 or 5, or deemed by the independent DMC to be serious enough to prevent an increase in dose of treatment.

### Secondary outcomes

Within 30 days of dosing:

1. Safety (assessed on days 0, 1, 2, 7 and 30)
  - Adverse events of particular interest (defined as transfusion reaction; macrophage activation syndrome; acute respiratory compromise) and all serious adverse events occurring within 30 days of infusion, clinical observations, clinical examination, electrocardiogram (ECG) and safety blood tests.
2. Activity (assessed on days 0, 1, 2, 7 and 30; if available from prior blood serum stored in the biochemistry laboratory, novel liver injury and inflammation markers at additional time points will also be included)
  - Pro-inflammatory – IL-6, TNF-alpha, IL-12, IL-8 (pg/mL). Anti-inflammatory - IL-10 (pg/mL). Assessed as change from baseline.
  - Liver injury – conventional markers of paracetamol-induced liver injury: alanine aminotransferase (U/L), International Normalised Ratio, Lactate (mmol/L), Creatinine (μmol/L). Novel marker of paracetamol-induced liver injury: High mobility group box 1 protein (ng/mL), glutamate dehydrogenase (U/L), cytokeratin-18 (U/L) and miR-122 (copies/mL). Assessed as change from baseline.
3. Immunogenicity (assessed on day 30)
  - Development of anti-human leukocyte antigen antibodies

## Recruitment

### Identification

Potentially eligible patients will be identified by their direct care team at the Emergency Department, Toxicology ward of RIE or from the Scottish Liver Transplantation Unit. An appropriately trained member of the Emergency Medicine Research Group nurses (EMERGE) will assess the patient for study eligibility. Appropriate participant information and informed consent forms will be provided (a copy is provided as Supplement 3). Consent will be obtained by an appropriately delegated investigator.

The recruitment period is expected to be two years, based on historic attendance numbers of potentially eligible patients.

### Randomisation

The dose assigned to the sentinel participant in each cohort will be fixed. For the remainder of cohorts 3, 4 and 5 randomisation, using computer-generated pseudo-random numbers produced by the programming team at Edinburgh Clinical Trials Unit (ECTU), will use the allocation ratios across available doses depicted in Figure 1a or 1b as appropriate.

### Allocation

A random permuted block will be used for each cohort to ensure the required allocation ratio is achieved. Allocation concealment will be maintained by use of a centralised online randomisation system.

### Blinding

In the randomised dose cohorts, the team that identify potential participants and screen for eligibility (EMERGE) will be blinded to the dose of cells delivered to a patient to minimise selection bias. EMERGE do not deliver any other trial activities post-baseline. The investigators may be involved in the consent process and baseline assessments but EMERGE will be solely responsible for identification and screening. Investigators, nurses in the RIECRF, who will be responsible for randomisation, dosing and follow-up, and the patient will know the allocated dose. The dose will not be documented in participants’ medical notes to reduce the likelihood of unblinding EMERGE. Blinding can end for individual participants when all participants of a particular dose cohort have been dosed; for instance, the recruiting team no longer need to be blinded for participants in dose cohort 3 when recruiting to dose cohort 4.

### Participant retention

If participants are not willing or able to return to the RIECRF, the day 7 and day 30 visits may be performed by a research nurse at the participant’s home. If participants are not willing to have an in-person visit at the RIECRF or at home, follow-up data will be collected by phone and/or using electronic health records. This will be explained to participants during the consenting process. If a participant does not return in person for their day 30 follow-up, this will not constitute a protocol deviation if information regarding clinical events, adverse events and concomitant medications can be collected by phone or from the patient’s medical records for the period between enrolment and the day 30 visit. On enrolment, participants will be provided with contact details for trial staff who can be contacted for advice. In cases of suspected adverse events, an unscheduled visit can be arranged to assess the participant.

## Study assessments

Study assessments are summarised in table 2. Research blood samples will be processed and labelled by research staff according to study-specific processing instructions, frozen at -80°C and then transferred periodically to the University of Edinburgh’s Centre for Regenerative Medicine on dry ice for storage in -80°C research freezers until analysis. Any prior blood serum samples received from the central RIE biochemistry laboratory at baseline will be stored and analysed in the same way.

**Table 2:**
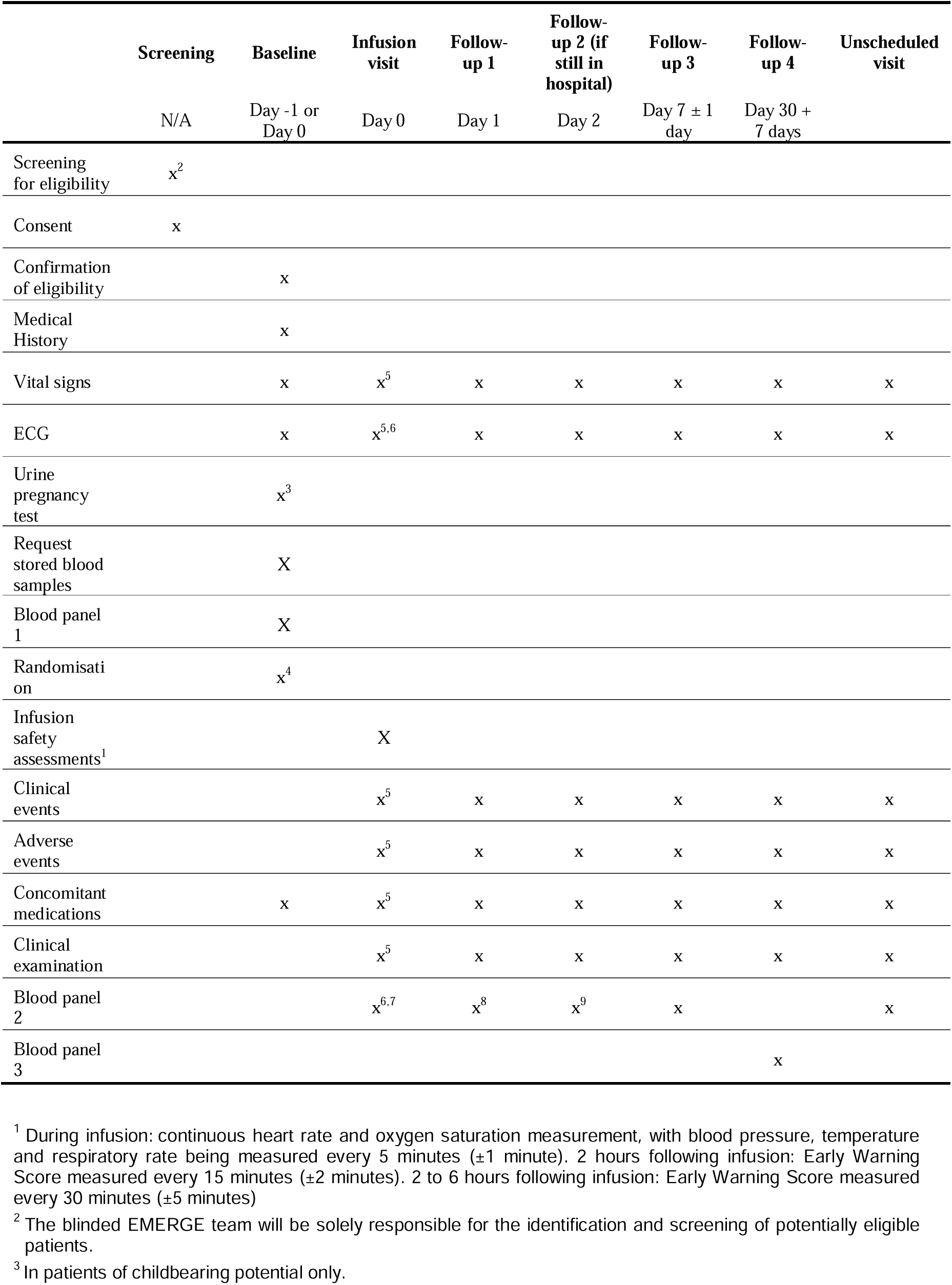
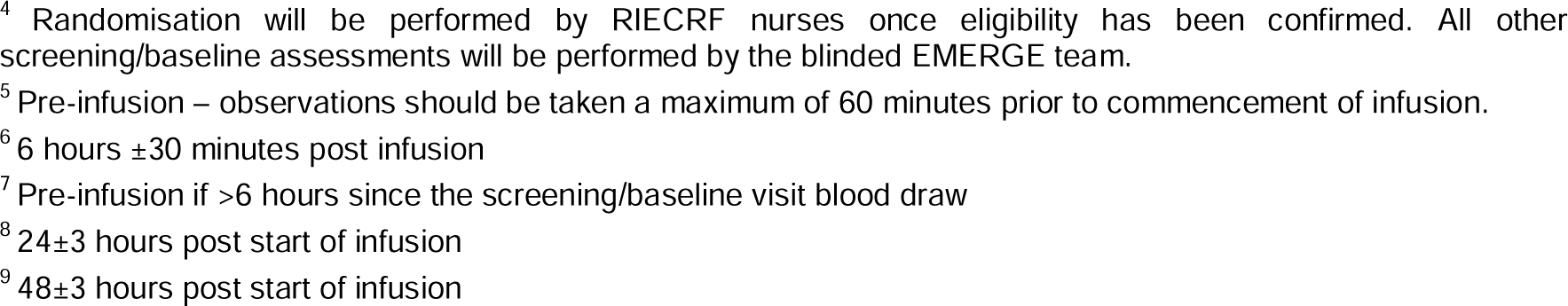
Study assessments.

|  | Screening | Baseline | Infusion visit | Follow-up 1 | Follow-up 2 (if still in hospital) | Follow-up 3 | Follow-up 4 | Unscheduled visit |
| --- | --- | --- | --- | --- | --- | --- | --- | --- |
|  | N/A | Day -1 or Day 0 | Day 0 | Day 1 | Day 2 | Day 7 ± 1 day | Day 30 + 7 days |  |
| Screening for eligibility | x<sup>2</sup> |  |  |  |  |  |  |  |
| Consent | x |  |  |  |  |  |  |  |
| Confirmation of eligibility |  | x |  |  |  |  |  |  |
| Medical History |  | x |  |  |  |  |  |  |
| Vital signs |  | x | x<sup>5</sup> | x | x | x | x | x |
| ECG |  | x | x<sup>5,6</sup> | x | x | x | x | x |
| Urine pregnancy test |  | x<sup>3</sup> |  |  |  |  |  |  |
| Request stored blood samples |  | X |  |  |  |  |  |  |
| Blood panel 1 |  | X |  |  |  |  |  |  |
| Randomisation |  | x<sup>4</sup> |  |  |  |  |  |  |
| Infusion safety assessments<sup>1</sup> |  |  | X |  |  |  |  |  |
| Clinical events |  |  | x<sup>5</sup> | x | x | x | x | x |
| Adverse events |  |  | x<sup>5</sup> | x | x | x | x | x |
| Concomitant medications |  | x | x<sup>5</sup> | x | x | x | x | x |
| Clinical examination |  |  | x<sup>5</sup> | x | x | x | x | x |
| Blood panel 2 |  |  | x<sup>6,7</sup> | x<sup>8</sup> | x<sup>9</sup> | x |  | x |
| Blood panel 3 |  |  |  |  |  |  | x |  |

<sup>1</sup> During infusion: continuous heart rate and oxygen saturation measurement, with blood pressure, temperature and respiratory rate being measured every 5 minutes (±1 minute). 2 hours following infusion: Early Warning Score measured every 15 minutes (±2 minutes). 2 to 6 hours following infusion: Early Warning Score measured every 30 minutes (±5 minutes)
<sup>2</sup> The blinded EMERGE team will be solely responsible for the identification and screening of potentially eligible patients.
<sup>3</sup> In patients of childbearing potential only.
<sup>4</sup> Randomisation will be performed by RIECRF nurses once eligibility has been confirmed. All other screening/baseline assessments will be performed by the blinded EMERGE team.
<sup>5</sup> Pre-infusion – observations should be taken a maximum of 60 minutes prior to commencement of infusion.
<sup>6</sup> 6 hours ± 30 minutes post infusion
<sup>7</sup> Pre-infusion if >6 hours since the screening/baseline visit blood draw
<sup>8</sup> 24 ± 3 hours post start of infusion
<sup>9</sup> 48 ± 3 hours post start of infusion

**Table 3:** Study blood tests.

|  | Blood panel 1 | Blood panel 2 | Blood panel 3 |
| --- | --- | --- | --- |
| Blood count | x | x | x |
| Clotting screen | x | x | x |
| Urea and electrolytes | x | x | x |
| Liver function tests | x | x | x |
| Triglycerides | x | x | x |
| Ferritin | x | x | x |
| Paracetamol | x | x | x |
| Research bloods (secondary outcomes) | x | x | x |
| C-reactive protein | x | x | x |
| Lactate | x | x | x |
| Liver antibodies | x |  |  |
| Immunoglobulins | x |  |  |
| Alpha fetoprotein | x |  |  |
| Alpha 1 antitrypsin | x |  |  |
| Caeruloplasmin | x |  |  |
| Calcium | x |  |  |
| Magnesium | x |  |  |
| Viral serology | x |  |  |
| HLA typing | x<sup>1</sup> |  |  |
| Anti-HLA antibodies | x<sup>2</sup> |  | x |

<sup>1</sup> HLA typing at baseline will be performed in all participants, with the result used to guide selection of donor AAMs.
<sup>2</sup> In patients with a history of pregnancy or blood product transfusion, this will be a rapid anti-HLA antibody screen. The result will be used to guide selection of donor AAMs.

## Use of study data

### Data collection

Study data can be recorded on paper data collection sheets and then entered into the electronic case report form (eCRF) by members of the research team at site. Alternatively, data may be entered directly into the eCRF. All electronic case report forms will be reviewed and approved by the Sponsor prior to use.

A copy of a blank eCRF can be requested by contacting.

### Data management

ECTU will provide and maintain a secure web-based REDCap database (Research Electronic Data Capture, Vanderbilt University, USA) compliant with Sponsor standard operating procedures. Trained delegated members of the study team will be given password-protected logins to the database for data entry. The data will be stored on a secure server in the University of Edinburgh for the minimum retention period for the study. No directly identifiable information will be entered into the database.

Authorised clients (such as the study or unblinded statistician needing access to produce progress reports to the independent DMC and/or interim or final statistical analyses; the Trial Manager to produce blinded logistics reports; and the Quality Assurance manager for audit and ongoing quality assurance reports) will be given protected access to relevant data.

### Data monitoring

A DMC, independent of the Sponsor and without competing interests, will be established to oversee the safety of participants in the trial. The terms of reference and composition of the DMC will be detailed in a charter in accordance with Sponsor requirements.

The DMC will review safety data and make dose progression recommendations for review by the Sponsor and Chief Investigator. They will also propose changes to the protocol and/or stop the study if indicated.

## Sample size and statistical analysis

If no safety concerns arise which lead to early stopping of the trial or modification of the dose escalation, the design outlined in Figure 1a will include 17 participants. If DLTs are observed at the 10^6^ and 10^7^ doses and the trial reverts to a standard 3+3 design, it will include up to 30 participants.

As well as assessment of the primary outcome of safety, this number of participants will enable exploration of any initial signals of efficacy on markers of paracetamol-induced liver injury such as HMGB1, through analysis by isotonic regression modelling. As a guide to the strengths of association detectable in this size of sample, in a one-way analysis of variance with 5 groups and a sample size of 3-4 per group, effect sizes (variance of means divided by within group standard deviation) between 1 and 2 can be detected with 80% power at a 5% significance level.

A detailed statistical analysis plan has been produced and prospectively documented within the Trial Master File in August 2023. The analysis population for safety outcomes will include all participants receiving any quantity of the investigational allogeneic macrophage cell therapy and will be analysed according to treatment received. Activity outcomes will be analysed for participants receiving any quantity of investigational AAM cell therapy according to intention to treat.

All DLT primary outcome events will be listed by dose and participant alongside their features of seriousness, intensity, relatedness, and outcome.

## Safety assessments

Patients will be in the RIECRF for a minimum of 6 hours after cell infusion. There will be continuous heart rate and oxygen saturation measurement during the cell infusion, with blood pressure, temperature and respiratory rate being measured every 5 minutes (±1 minute). The CI or an appropriately trained and delegated sub-investigator, and the RIECRF nurse will be physically present with the patient for the duration of the cell infusion and for 2 hours after dosing with the Early Warning Score (EWS) measured every 15 minutes (±2 minutes). From 2 to 6 hours after dosing, the EWS will be measured every 30 minutes (±5 minutes), with the RIECRF nurse being physically present with the patient, and the CI or an appropriately trained and delegated sub-investigator being present on site.

Participant will remain in hospital overnight after dosing. There will be a handover of information from the research team to the clinical team and the Principal Investigator or an appropriately trained and delegated sub-investigator will be contactable overnight. The patient will be assessed the following day by the research team. Discharge will be a clinical decision of the caring consultant/team.

There are some safety outcomes of particular interest, which have not been reported in previous human trials of macrophage therapy but remain theoretical risks (Table 4).

**Table 4.** Safety outcomes of particular interest and the criteria against which they will be assessed.

| Outcome | Criteria |
| --- | --- |
| Acute transfusion reaction | • Temperature $\geq 38^{\circ}\text{C}$ or raise of $2^{\circ}\text{C}$<br>• HR $> 100/\text{minute}$<br>• Fall in blood pressure of $\geq 30\text{mmHg}$<br>• Collapse<br>• Pain (chest or abdominal or along infusion site)<br>• Dyspnoea or oxygen saturation $< 94\%$ in room air<br>• Stridor<br>• Mucosal swelling<br>• New rash<br>• Bleeding diathesis<br>• Any other significant changes in clinical condition or observation. |
| Macrophage activation syndrome | 1. Fever<br>2. Cytopenia in 2 or more lineages<br>i. Hb $< 10.0\text{ g/dL}$<br>ii. Platelets $< 100 \times 10^9/\text{L}$<br>iii. Neutrophils $< 1.0 \times 10^9/\text{L}$<br>3. Hypertriglyceridemia or hypofibrinogenemia<br>i. Triglycerides $> 2.99\text{ mmol/L}$<br>ii. Fibrinogen $< 1.50\text{ g/L}$<br>4. Ferritin $> 500\text{ }\mu\text{g/L}$ |
| Acute respiratory compromise | • SpO<sub>2</sub> $< 92\%$ on air, or need for supplemental oxygen<br>• Increase in respiratory rate $> 20/\text{minute}$ |

## Monitoring and oversight

All patient enrolment will stop pending advice from the DMC and a representative of the Sponsor if one of the following stopping rules have been met:

- a patient dies or requires an escalation in their clinical care with admission to the critical care unit or liver transplant unit after receiving the trial treatment
- a patient has a safety outcome of particular interest – transfusion reaction, macrophage activation syndrome or acute respiratory compromise as detailed in table 4.
- a suspected unexpected serious adverse reaction occurs.

All clinical events occurring in this study will be assessed against these criteria by an investigator within 24 hours of discovery. If a stopping rule has been met, the investigator will notify the trial team and Sponsor within 24 hours of discovery. The trial management team will communicate the recruitment stop to all relevant parties and arrange DMC review.

Following the single infusion of AAMs and beyond the end of the trial, participants will receive standard clinical care. NHS indemnity scheme will apply and the University has No Fault Compensation Insurance in place.

## Study amendments and current status

At the time of submission, the protocol in operation is version 5.0, and this is reflected in this manuscript. Protocol changes and implementation dates are provided as Supplement 1. Recruitment started on September 2023. The trial is following scenario 1 and the study team have recruited the first participant in Cohort 3. We are on target to complete recruitment by the end of 2025.

Participants 1 and 2 were recruited under protocol version 3.0. Participant 3 was recruited under protocol version 5.0. The protocol changes have served only to clarify processes and would not have impacted participant eligibility.

Any future amendments to the protocol will be communicated to all relevant parties, including trial registries and publishing journals.

## Discussion

MAIL is a randomised trial designed to determine whether there is DLT of allogeneic AAMs in patients with paracetamol-induced ALI, and assess the activity of AAMs across a range of secondary outcomes. It builds on previous macrophage therapy work to identify a treatment for a patient group who currently experience high morbidity and mortality.

Pre-clinical studies in mice have shown promise, and importantly, efficacy even in established liver injury.(12) The therapy is deliverable by peripheral venous access, and has the potential to be delivered in any acute care setting which delivers blood products.

Previous trials have focused on autologous cell therapies, which due to production lead times and decreased monocyte counts is not an option for patients with paracetamol-induced ALI.(13,18) This trial has the potential to provide evidence on the safety and tolerability of an ‘off-the-shelf’ cell therapy for patients in unscheduled care.

This trial will potentially lay the groundwork for a phase 2 trial of AAMs in paracetamol-induced ALI; the additional data collected during MAIL will inform the design of that future trial.

## Ethics and dissemination

The trial will be conducted according to the ethical principles of the Declaration of Helsinki 2013 and has been approved by North East – York Research Ethics Committee (reference 23/NE/0019), NHS Lothian Research and Development department, and the MHRA. Good Clinical Practice regulations will be followed and written informed consent will be obtained from all participants.

No trial results will be shared while the trial is ongoing. When the trial completes, data will be shared via presentation, publication and may be available on request from the trial Chief Investigator, decided on a case-by-case basis. Data sharing may be restricted in line with patent or commercial requirements for as short a time as possible.

Results will be disseminated by peer-reviewed publication, conferences, and linked on isrctn.com. Publication content will follow the CONSORT-DEFINE reporting guidance extension for dose-finding studies.(19) Ownership of data arising from this study resides with the study team and their respective employers. The study team will follow the International Committee of Medical Journal Editors guidelines on authorship. Requests for data access should be sent to the corresponding author.

## Funding

This work is supported by the Medical Research Council (MRC) [MR/T044802/1]; and The Centre for Precision Cell Therapy for the Liver which was funded by the Chief Scientist Office (CSO) of the Scottish Government Health Directorates [PMAS/21/07]. For the purpose of open access, the author has applied a creative commons attribution (CC BY) licence to any author accepted manuscript version arising.

## Authorship statement

All of the MAIL Trial Investigators authorship group made substantial contributions to the conception or design of the work; and either drafting the work or reviewing it critically for important intellectual content; and final approval of the version to be published; and all agree to be accountable for all aspects of the work in ensuring that questions related to the accuracy or integrity of any part of the work are appropriately investigated and resolved.

## Declaration of interests

All authors acknowledge that work for the MAIL Trial and publication of this manuscript is supported by MRC and CSO funding. GA: consulting fees from Clinipace, GSK, Amryth, Pfizer, DNDi, BenevolentAI Bio, PureTech LYT, Merck, Agios, Astra Zeneca, Suzhou MDCE. JCa: advisor to, founding member of, and stock options in Resolution Therapeutics, multiple patents related to delivery of macrophage cell therapy. JD: MRC funding (and patent filed) for an in vitro diagnostic which could be a companion diagnostic, CSO funding for another clinical trial for treatment of paracetamol overdose, scientific advisory board member for EU funded TransBioLine consortium. RF: grants or contracts from Takeda and Kezar. SF: consulting fees for Cytotheryx and Resolution Therapeutics, honoraria received from Japanese Society for Regenerative Medicine, multiple patents related to macrophage cell therapy, advisory board member for Cytotheryx, stock options, founder, and director of Resolution Therapeutics. CH: grants from the Centre for Precision Cell Therapy for the Liver, honoraria from Elsevier, support for meetings and travel from Royal College of Emergency Medicine (RCEM), member of RCEM Toxicology advisory group, editor for a BMJ Group medical journal. ARo: independent statistician for other clinical trials.

## Supporting information

SPIRIT-DEFINE

## Data Availability

Results will be disseminated by peer-reviewed publication, conferences, and linked on isrctn.com. Publication content will follow the CONSORT-DEFINE reporting guidance extension for dose-finding studies. Ownership of data arising from this study resides with the study team and their respective employers. The study team will follow the International Committee of Medical Journal Editors guidelines on authorship. Requests for data access should be sent to the corresponding author.

## Glossary

AAMs: Alternatively Activated Macrophages
ALF: Acute Liver Failure
ALI: Acute Liver Injury
ATMP: Advanced Therapy Medicinal Product
CTIMP: Clinical Trial of Investigational Medicinal Product
DLT: Dose-Limiting Toxicity
DMC: Data Monitoring Committee
eCRF: Electronic Case Report Form
ECTU: Edinburgh Clinical Trials Unit
EMERGE: Emergency Medicine Research Group of Edinburgh
HLA: Human Leukocyte Antigen
MHRA: United Kingdom Medicines and Healthcare products Regulatory Agency
NAC: Acetylcysteine
POD: Paracetamol (acetaminophen) overdose
RIECRF: Royal Infirmary of Edinburgh Clinical Research Facility
TSC: Trial Steering Committee

## Notes

### Clinical Trial

ISRCTN12637839

### Author Declarations

The trial will be conducted according to the ethical principles of the Declaration of Helsinki 2013 and has been approved by North East - York Research Ethics Committee (reference 23/NE/0019), NHS Lothian Research and Development department, and the MHRA. Good Clinical Practice regulations will be followed and written informed consent will be obtained from all participants.

