## Supplementary material for "Macrophage Therapy for Acute Liver Injury (MAIL): a Phase 1 Randomised, Open-Label, Dose-Escalation Study to Evaluate Safety, Tolerability, and Activity of Allogeneic Alternatively Activated Macrophages in Patients with Paracetamol-induced Acute Liver Injury": SPIRIT-DEFINE

### Web appendix 2: SPIRIT-DEFINE downloadable checklist

**Recommended checklist items to address in an early phase dose-finding (EPDF) clinical trial protocol from SPIRIT 2013 and SPIRIT-DEFINE checklists^**

Please cite as: Yap C, Rekowski J, Ursino M, et al. Enhancing quality and impact of early phase dose-finding clinical trial protocols: SPIRIT Dose-finding Extension (SPIRIT-DEFINE) guidance. *BMJ* 2023;383;e076836. doi:10.1136/bmj-2023-076386

| **Category and section** | **Standard SPIRIT 2013 checklist item** | | **SPIRIT-DEFINE checklist item**  **for EPDF trials** | | **Addressed on Page No^¤^** |
| --- | --- | --- | --- | --- | --- |
|  | **Item No** | **SPIRIT 2013** | **Item No** | **SPIRIT-DEFINE** |  |
| **Administrative information** | | | | | |
| Title | 1 | Descriptive title identifying the study design, population, interventions, and, if applicable, trial acronym | 1† | Descriptive title identifying the early phase dose-finding trial design (eg, dose escalation or de-escalation, placebo controlled, multiple ascending dose), population, interventions, and whether the trial was randomised, and, if applicable, trial acronym | 1 |
| Trial registration | 2a | Trial identifier and registry name. If not yet registered, name of intended registry | 2a |  | 1 |
|  | 2b | All items from the World Health Organization Trial Registration Data Set | 2b |  | 1-16 |
| Protocol version | 3 | Date and version identifier | 3 |  | 1 |
| Funding | 4 | Sources and types of financial, material, and other support | 4 |  | 6,15 |
| Roles and responsibilities | 5a | Names, affiliations, and roles of protocol contributors | 5a |  | 1 |
|  | 5b | Name and contact information for the trial sponsor | 5b |  | 6 |
|  | 5c | Role of study sponsor and funders, if any, in study design; collection, management, analysis, and interpretation of data; writing of the report; and the decision to submit the report for publication, including whether they will have ultimate authority over any of these activities | 5c |  | 6 |
|  | 5d | Composition, roles, and responsibilities of the coordinating centre, steering committee, endpoint adjudication committee, data management team, and other individuals or groups overseeing the trial, if applicable (see item 21a for data monitoring committee) | 5d |  | Supplement |
| **Introduction** | | | | | |
| Background and rationale | 6a | Description of research question and justification for undertaking the trial, including summary of relevant studies (published and unpublished) examining benefits and harms for each intervention | 6a.1† | Description of research question(s) and justification for undertaking the trial, including summary of relevant clinical studies (published and unpublished) examining benefits and harms for each intervention | 2 |
|  |  |  | 6a.2* | Summary of key findings from relevant non-clinical or preclinical research | 2,3 |
|  |  |  | 6a.3* | Summary of findings from previously generated preclinical and translational studies to support any planned biomarker substudies (where applicable) | 2,3 |
|  | 6b | Explanation for choice of comparators | 6b |  | N/A |
| Objectives | 7 | Specific objectives or hypotheses | 7† | Specific objectives (eg, relating to safety, activity, pharmacokinetics, pharmacodynamics, recommended dose(s)) | 3 |
| Trial design | 8 | Description of trial design including type of trial (eg, parallel group, crossover, factorial, single group), allocation ratio, and framework (eg, superiority, equivalence, noninferiority, exploratory) | 8a.1† | Description of trial design elements, such as dose escalation or de-escalation strategy, number of treatment groups, allocation ratio if relevant, and details of any prespecified trial adaptations | 3-6 |
|  |  |  | 8a.2* | Trial design schema to show the flow of major transition points (eg, dose escalation to dose expansion, phase 1 to phase 2, single ascending dose to multiple ascending dose) | 4-6 |
|  |  |  | 8a.3* | Statistical methods or rationale underpinning the trial design | 3-6 |
|  |  |  | 8a.4* | Prespecified interim decision making criteria or rules to guide the trial adaptation process (eg, dose escalation or de-escalation, early stopping, progression to the next part of the trial); planned timing and frequency of interim data looks and the information to inform the adaptations; alternatively, an explanation of why they are not prespecified | 3-6 |
|  |  |  | 8a.5* | Starting dose(s) with rationale | 3 |
|  |  |  | 8a.6* | Range of planned dose levels with rationale | 3 |
|  |  |  | 8a.7* | Presentation of planned dose levels (eg, as a diagram, table, or infographic), where applicable | 4-6 |
|  |  |  | 8a.8* | Skipping of dose level(s), if applicable | N/A |
|  |  |  | 8a.9* | Planned cohort size(s) (eg, fixed, flexible, adaptive) | 3-6 |
|  |  |  | 8a.10* | Dose allocation method within a dose level (including sequence and interval between dosing of participants, eg, sentinel or staggered dosing) | 3-6 |
|  |  |  | 8a.11* | Dose expansion cohort(s), if applicable, with rationale | 3-6 |
| **Methods: Participants, interventions, and outcomes** | | | | | |
| Study settings | 9 | Description of study settings (eg, community clinic, academic hospital) and list of countries where data will be collected. Reference to where list of study sites can be obtained | 9 |  | 6 |
| Eligibility criteria | 10 | Inclusion and exclusion criteria for participants. If applicable, eligibility criteria for study centres and individuals who will perform the interventions (eg, surgeons, psychotherapists) | 10 |  | 7,8 |
| Interventions | 11a | Interventions for each group with sufficient detail to allow replication, including how and when they will be administered | 11a† | Interventions for each dose level (within each group) with sufficient details to allow replication, including administration route and schedule showing how and when they will be administered | 8,9 |
|  | 11b | Criteria for discontinuing or modifying allocated interventions for a given trial participant (eg, drug dose change in response to harms, participant request, or improving/worsening disease) | 11b† | Criteria for dose discontinuation, dose modifications, and dosing delays of allocated interventions for a given trial participant (eg, dose change in response to harms, participant request, or improving or worsening disease) | 9 |
|  | 11c | Strategies to improve adherence to intervention protocols, and any procedures for monitoring adherence (eg, drug tablet return, laboratory tests) | 11c |  | 10 |
|  | 11d | Relevant concomitant care and interventions that are permitted or prohibited during the trial | 11d |  | 3,9 |
| Outcomes | 12 | Primary, secondary, and other outcomes, including the specific measurement variable (eg, systolic blood pressure), analysis metric (eg, change from baseline, final value, time to event), method of aggregation (eg, median, proportion), and time point for each outcome. Explanation of the clinical relevance of chosen efficacy and harm outcomes is strongly recommended | 12† | Primary, secondary, and other outcomes (which include those intended for prespecified adaptations), including the specific measurement variable, analysis metric, method of aggregation, and time point for each outcome. Explanation of the clinical relevance of chosen outcomes is strongly recommended. | 9-12 |
| Participant timeline | 13 | Time schedule of enrolment, interventions (including any run-ins and washouts), assessments, and visits for participants. A schematic diagram is highly recommended | 13† | Time schedule of enrolment, interventions (including any run-ins and washouts), assessments, and visits for participants (including in-house stay or out-patient follow-up period, if applicable); a schematic diagram is highly recommended | 7 |
| Sample size | 14 | Estimated number of participants needed to achieve study objectives and how it was determined, including clinical and statistical assumptions supporting any sample size calculations | 14† | Estimated number of participants (minimum, maximum, or expected range) needed to address trial objectives and how it was determined, including clinical and statistical assumptions supporting any sample size and operating characteristics | 12 |
| Recruitment | 15 | Strategies for achieving adequate participant enrolment to reach target sample size | 15 |  | 10 |
| **Methods: Assignment of interventions (for controlled trials)** | | | | | |
| Allocation: Sequence generation | 16a | Method of generating the allocation sequence (eg, computer generated random numbers), and list of any factors for stratification. To reduce predictability of a random sequence, details of any planned restriction (eg, blocking) should be provided in a separate document that is unavailable to those who enrol participants or assign interventions | 16a.1 |  | 10 |
|  |  |  | 16a.2* | Any prespecified rule or algorithm to update allocation with timing and frequency of updates, if applicable | 10 |
| Allocation concealment mechanism | 16b | Mechanism of implementing the allocation sequence (eg, central telephone; sequentially numbered, opaque, sealed envelopes), describing any steps to conceal the sequence until interventions are assigned | 16b |  | 10 |
| Implementation | 16c | Who will generate the allocation sequence, who will enrol participants, and who will assign participants to interventions | 16c |  | 10 |
| Blinding (masking) | 17a | Who will be blinded after assignment to interventions (eg, trial participants, care providers, outcome assessors, data analysts), and how | 17a |  | 10 |
|  | 17b | If blinded, circumstances under which unblinding is permissible, and procedure for revealing a participant’s allocated intervention during the trial | 17b |  | 3,10 |
| **Methods: Data collection, management, and analysis** | | | | | |
| Data collection methods | 18a | Plans for assessment and collection of outcome, baseline, and other trial data, including any related processes to promote data quality (eg, duplicate measurements, training of assessors) and a description of study instruments (eg, questionnaires, laboratory tests) along with their reliability and validity, if known. Reference to where data collection forms can be found, if not in the protocol | 18a |  | 11,12 |
|  | 18b | Plans to promote participant retention and complete follow-up, including list of any outcome data to be collected for participants who discontinue or deviate from intervention protocols | 18b |  | 10 |
| Data management | 19 | Plans for data entry, coding, security, and storage, including any related processes to promote data quality (eg, double data entry; range checks for data values). Reference to where details of data management procedures can be found, if not in the protocol | 19 |  | 12 |
| Statistical methods | 20a | Statistical methods for analysing primary and secondary outcomes. Reference to where other details of the statistical analysis plan can be found, if not in the protocol | 20a.1† | Statistical methods for primary and secondary outcomes and any other outcomes used to make prespecified adaptations; reference to where other details of the statistical analysis plan can be accessed, if not in the protocol | 12 |
|  |  |  | 20a.2* | For the proposed adaptive design features, statistical methods used for estimation (eg, safety, dose(s), treatment effects) and to make inferences | 13 |
|  | 20b | Methods for any additional analyses (eg, subgroup and adjusted analyses) | 20b† | Statistical methods for additional analyses (eg, subgroup and adjusted analyses, pharmacokinetics or pharmacodynamics, biomarker correlative analyses) | N/A |
|  | 20c | Definition of analysis population relating to protocol non-adherence (eg, as randomised analysis), and any statistical methods to handle missing data (eg, multiple imputation) | 20c.1† | Analysis population(s) (eg, evaluable population for dose-finding, safety population) | 13 |
|  |  |  | 20c.2* | Strategies for handling intercurrent events occurring after treatment initiation (eg, how dosing adjustments will be handled) that can affect either the interpretation or the existence of the measurements associated with the clinical question of interest, and any methods to handle missing data | 12 |
| **Methods: Data monitoring** | | | | | |
| Data monitoring – formal committee | 21a | Composition of DMC; summary of its role and reporting structure; statement of whether it is independent from the sponsor and competing interests; and reference to where further details about its charter can be found, if not in the protocol. Alternatively, an explanation of why a DMC is not needed | 21a† | Composition of any decision making or safety review committee or group; summary of its role and reporting structure; statement of whether it is independent from the sponsor and competing interests; and reference to where further details, such as a charter, can be found, if not in the protocol; alternatively, an explanation of why such a committee is not needed | Supplement |
| Data monitoring – interim analyses | 21b | Description of any interim analyses and stopping guidelines, including who will have access to these interim results and make the final decision to terminate the trial | 21b† | Description of who will have access to interim results and make the interim and final decision to terminate the trial (or part(s) of the trial, eg, end of dose escalation), and measures to safeguard the confidentiality of interim information | 13 |
| Harms | 22 | Plans for collecting, assessing, reporting, and managing solicited and spontaneously reported adverse events and other unintended effects of trial interventions or trial conduct | 22† | Plans for collecting, assessing, reporting, and managing solicited and spontaneously reported harms such as adverse events (eg, toxicities) and other unintended effects of trial interventions or trial conduct, including time frames of reporting these events or effects to allow informed interim decision making (eg, before any planned next dosing) | 13 |
| Auditing | 23 | Frequency and procedures for auditing trial conduct, if any, and whether the process will be independent from investigators and the sponsor | 23 |  | 12 |
| **Ethics and dissemination** | | | | | |
| Research ethics approval | 24 | Plans for seeking REC/IRB approval | 24 |  | 14 |
| Protocol amendments | 25 | Plans for communicating important protocol modifications (eg, changes to eligibility criteria, outcomes, analyses) to relevant parties (eg, investigators, REC/IRBs, trial participants, trial registries, journals, regulators) | 25 |  | 15 |
| Consent or assent | 26a | Who will obtain informed consent or assent from potential trial participants or authorised surrogates, and how (see item 32) | 26a |  | 10 |
|  | 26b | Additional consent provisions for collection and use of participant data and biological specimens in ancillary studies, if applicable | 26b |  | 10 |
| Confidentiality | 27 | How personal information about potential and enrolled participants will be collected, shared, and maintained in order to protect confidentiality before, during, and after the trial | 27 |  | 12 |
| Declaration of interests | 28 | Financial and other competing interests for principal investigators for the overall trial and each study site | 28 |  | 17 |
| Access to data | 29 | Statement of who will have access to the final trial dataset, and disclosure of contractual agreements that limit such access for investigators | 29 |  | 12 |
| Ancillary and post-trial care | 30 | Provisions, if any, for ancillary and post-trial care, and for compensation to those who experience harm from trial participation | 30 |  | 14 |
| Dissemination policy | 31a | Plans for investigators and sponsor to communicate trial results to participants, healthcare professionals, the public, and other relevant groups (eg, via publication, reporting in results databases, or other data sharing arrangements), including any publication restrictions | 31a.1 |  | 14 |
|  |  |  | 31a.2* | Plans for sharing results (eg, safety, activity) externally while the trial is still ongoing, if applicable | 15 |
|  | 31b | Authorship eligibility guidelines and any intended use of professional writers | 31b |  | 15 |
|  | 31c | Plans, if any, for granting public access to the full protocol, participant level dataset, and statistical code | 31c |  | N/A |
| Informed consent materials | 32 | Model consent form and other related documentation given to participants and authorised surrogates | 32 |  | Supplement |
| Biological specimens | 33 | Plans for collection, laboratory evaluation, and storage of biological specimens for genetic or molecular analysis in the current trial and for future use in ancillary studies, if applicable | 33 |  | 11 |
| **Appendices** | | | | | |
| Dose transition pathways |  |  | 34* | Dose transition pathways or dose decision paths (using, eg, a flow diagram or table) projecting in advance how a proposed dose-finding design will recommend doses based on participants’ key outcomes | 3-6 |

DEFINE=Dose-finding Extension; DMC=data monitoring committee; EPDF=early phase dose-finding IRB=institutional review board; REC=research ethics committee; SPIRIT=Standard Protocol Items: Recommendations for Interventional Trials.

^^^ This checklist should be read in conjunction with the SPIRIT 2013 Explanation & Elaboration (1) for important clarification on the items. Amendments to the protocol should be tracked and dated. Empty items in the SPIRIT-DEFINE column indicate no modification from the SPIRIT 2013 items.

¤ If the information is not provided in the primary paper, give details of where this information is available. This may include locations such as published papers (provide citation details) or a website (provide the URL).

* New items that should only be applied in reference to SPIRIT-DEFINE.

† Modified items that require reference to both SPIRIT 2013 and SPIRIT-DEFINE.

The term “dose” in the checklist might be considered synonymous and used interchangeably with dosage or dosing regimen (dose and schedule) or a unit dose.

The SPIRIT checklist is copyrighted by the SPIRIT Group under the Creative Commons “[Attribution-NonCommercial-NoDerivs 3.0 Unported](http://www.creativecommons.org/licenses/by-nc-nd/3.0/)” license and reproduced with permission.
