## Supplement 1 for "Macrophage Therapy for Acute Liver Injury (MAIL): a Phase 1 Randomised, Open-Label, Dose-Escalation Study to Evaluate Safety, Tolerability, and Activity of Allogeneic Alternatively Activated Macrophages in Patients with Paracetamol-induced Acute Liver Injury"

| **Protocol Version** | **Implementation date** | **Changes** | **Comment** |
| --- | --- | --- | --- |
| V1.0 12Dec2022 | NA | NA | Submitted with the initial application to REC and MHRA but never approved |
| V2.0 27Feb2023 | NA | - Section 6.1.4 amended to clarify where IMP  labelling and QP release will take place - Definition of sexual abstinence added to section 4.2 | Submitted in response to request for further information; first version approved by REC and MHRA |
| V3.0 11May2023 | 01Sep2023 | - Table 4 Blood tests - measurement of cytokeratin-18 from fingerprick capillary sample removed as cytokeratin-18 will be measured in research blood panel - Section 7.2 Study assessments - minor clarifications/corrections (concomitant medication check included at baseline, clinical exam includes neurological system, maximum/minimum of any routine clinical vital sign measurements recorded at follow-ups 1 and 2) - Section 7.5 Blood sample analysis - 2.7mL fluoride ECTA tube added for measurement of lactate (previously omitted by mistake) and samples collected for HLA typing and antibody testing clarified - Section 11 Pharmacovigilance - minor additions to make it clear it is a first in human trial - REC and ISRCTN reference numbers added - Typographical errors corrected throughout - Minor clarifications to various sections | Submitted as part of non-substantial amendment 11May2023. First version implemented when site opened |
| V4.0 22Jan2024 | NA | - Inclusion criterion 1 (section 4.2) was amended to clarify the timing of serum ALT activity >1000U/L. Section 5.4 was updated to include further clarification on this. - Exclusion criterion 3 (section 4.3) was amended to clarify that current infections are excluded only. - Exclusion criterion 7 (section 4.3) was amended to exclude patients that previously received any organ transplant. - Section 5.1 was updated to clarify how potentially eligible patients are being identified. - Section 5.6.3 was updated to clarify what activities are being completed by the blinded EMERGE team. The previously used term 'recruiting team' was updated throughout to avoid confusion. - Section 6.1.1 was amended to clarify how HLA testing at baseline is used to guide product selection. A footnote was added to Table 4 in section 7.2 for the same purpose. - Sections 6.3.2 and 6.7.1 were updated to provide a more specific time window for the administration of pre-medication. - Section 7 was updated to differentiate more clearly between screening and baseline activities. Figure 1 in section 3 was updated in accordance with this. - Section 7.2 was amended to clarify that up to five prior blood serum samples will be requested from the laboratory (rather than all available samples). - CTCAE grade 5 was added to the definition of Dose Limiting Toxicity throughout (previously omitted in error). - Reference to specific training in assessing capacity was removed from section 5.2 (previously included in error). - In section 7.2, day 7 was included in the statement regarding follow-up data collection and deviations (previously omitted in error). - Reference to MedDRA coding of Adverse Events was removed from section 11.5 (previously included in error). | Submitted as part of non-substantial amendment 25Jan2024 but never implemented as superseded by protocol v5.0. |
| V5.0 07Mar2024 | 13Mar2024 | - The changes to protocol version 4.0 (listed above) were incorporated into V5.0 - Minor correction in Table 4, clarifying that the serology blood tests performed for Hepatitis B include a total 'core antibody' test rather than a  'core IgM' test. | Current version |
