## Supplement 2 for "Macrophage Therapy for Acute Liver Injury (MAIL): a Phase 1 Randomised, Open-Label, Dose-Escalation Study to Evaluate Safety, Tolerability, and Activity of Allogeneic Alternatively Activated Macrophages in Patients with Paracetamol-induced Acute Liver Injury"

### Supplemental information – committee compositions and roles

#### Trial Steering Committee

The TSC has six members, the Chief Investigator, the Trial Manager, a Lay member and three individuals with expertise in this clinical area. The TSC will provide oversight for the trial on behalf of the Sponsor/funder. The specific roles of the TSC include; monitoring recruitment rates and encouraging the Steering Group (SG) to develop strategies to deal with recruitment issues, review regular reports of the trial and assess the impact and relevance of any accumulating external evidence. It should also provide advice through its independent Chair to the SG on all aspects of the trial. The TSC will be responsible for reviewing the DMC recommendations, to determine whether amendments to the protocol or changes in study conduct are required.

#### Steering Group

The SG consists of the Principal Investigator, Chief and Co-investigators and individuals from the research team, trial management team and the Scottish National Blood Transfusion Service. Their role is to oversee the day to day running of the study and implement recommendations from the other committees.

#### Regulatory and Clinical Trial Group

This committee consists of the Chief and Co-investigators, individuals from the research team, trial management team, the statistician and SNBTS. Their role is to oversee the running of the study and the manufacture and supply of the clinical product.

#### Data Monitoring Committee

The DMC is an independent multidisciplinary group consisting of four clinicians and statisticians that, collectively, have experience/expertise in the management of patients with Acute Liver Injury and anticipated adverse effects and in the conduct and monitoring of clinical trials. All four members of the DMC are independent from the sponsor and have no competing interests.

The DMC will be responsible for safeguarding the interests of trial participants, potential participants, investigators and sponsor; assessing the safety and activity of the intervention during the trial; reviewing external evidence with an impact on risk/benefit balance and for monitoring the overall conduct of the clinical trial. The DMC will provide recommendations about stopping, modifying or continuing the trial to the Chief Investigator, Sponsor and Trial Steering Committee. To contribute to enhancing the integrity of the trial, the DMC may also formulate recommendations relating to the selection, recruitment, or retention of participants, or their management, or to improving their adherence to protocol-specified regimens and retention of participants, and the procedures for data management and quality control.
