## Supplement 3 for "Macrophage Therapy for Acute Liver Injury (MAIL): a Phase 1 Randomised, Open-Label, Dose-Escalation Study to Evaluate Safety, Tolerability, and Activity of Allogeneic Alternatively Activated Macrophages in Patients with Paracetamol-induced Acute Liver Injury"

**Participant Information Sheet**

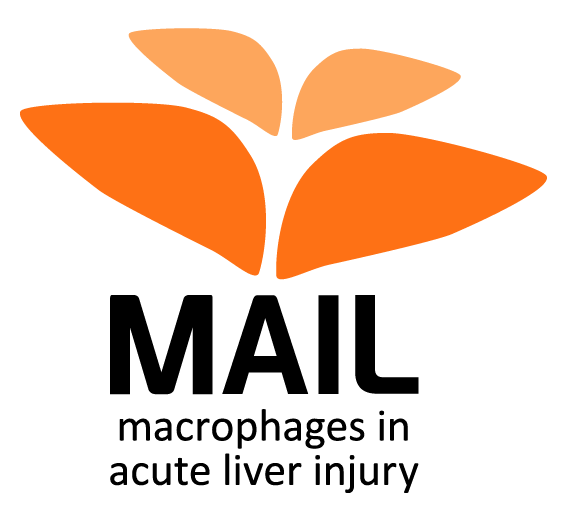

**You are invited to take part in a research study. To help you decide whether or not to take part, it is important for you to understand why the research is being done and what it will involve. Please take time to read the following information carefully. Talk to others about the study if you wish. Contact us if there is anything that is not clear, or if you would like more information. Take time to decide whether or not you wish to take part.**

**What is the purpose of the study?**

As you may know, paracetamol can be harmful to the liver if too much is taken. To help prevent liver damage, a drip with a medicine called acetylcysteine is given. However, some patients present to hospital and acetylcysteine is not effective because too much time has passed since the paracetamol was taken. Previous research has shown that macrophages (large white blood cells which have the ability to clear away damaged cells) can help to reduce the damage and support regeneration of the liver. This study will test whether a new treatment using macrophages is safe and can be beneficial for patients with liver damage caused by a paracetamol overdose. We hope to recruit up to 30 patients to this study.

**Why have I been invited to take part?**

We are inviting you to take part in this study because paracetamol has caused a degree of damage to your liver.

**Do I have to take part?**

No, it is up to you to decide whether or not to take part. If you do decide to take part you will be given this information sheet to keep and be asked to sign a consent form. If you decide to take part you are still free to withdraw at any time and without giving a reason. Deciding not to take part or withdrawing from the study will not affect the healthcare that you receive, or your legal rights.

**What will happen if I take part?**

A trained member of the research team will speak to you to discuss this study and to make sure you understand everything. We will give you time to ask questions and to decide if you want to take part. The amount of time will depend on how quickly you require treatment, but we will give you at least 1 hour. You will then be asked to sign the consent form.

If you decide to take part, we will carry out different tests to make sure it is safe for you to go into the study. Some of the blood you have given will be tested for certain infections (including Hepatitis A, B, C and E and human immunodeficiency virus (HIV), Cytomegalovirus and Epstein-Barr Virus. Because of your liver condition, you may have already been tested for some of these infections. The results of these tests will be discussed with you and if necessary, we will arrange for you to see an appropriate specialist. We will ask you about your medical history and any other medications you take, and make routine checks of your general well-being such as taking your temperature, blood pressure, breathing and pulse rate; and oxygen levels in your blood. We will also perform a tracing of the heart called an electrocardiogram (ECG). We will also take 11 tubes of blood for tests (approximately 8 teaspoons).

Treatment (Day 0)

If you are found to be suitable for the study, we will arrange for you to receive the study treatment. This could happen either on the same day as the tests described above or on the following day. You will be transferred from the ward to the Royal Infirmary of Edinburgh Clinical Research Facility, also in the Royal Infirmary of Edinburgh. Before you receive the treatment, a doctor will do a physical examination and we will repeat the routine checks of your general well-being and the ECG. We will also ask you about any other medications you take and any side effects you are experiencing from the paracetamol overdose or any other treatment you received in hospital. If it has been longer than 6 hours since we took blood samples from you, we will take blood again (approximately 4 teaspoons).

For the treatment, we will place a needle into one of your arms. Firstly, we will give you an antihistamine through this needle to reduce the likelihood of an allergic reaction. The needle will then be connected to a bag (via a plastic tube) which will contain the blood cells for infusion. The dose of cells you receive will depend on how many people have already taken part in the study as the dose of cells slowly increases as the study goes on. You will be observed at the Royal Infirmary of Edinburgh Clinical Research Facility during the infusion and for a minimum of 6 hours after the infusion, with checks of your general well-being being performed approximately every 15 minutes for the first 2 hours and every 30 minutes for the next 4 hours. Around 6 hours after the infusion, we will take 6 tubes of blood for tests (approximately 4 teaspoons), perform an ECG and then transfer you back to the ward.

We will give you a small card with information about the trial and emergency contact details. Please carry this card with you while you are participating in the study. If you see a doctor, please show it to them.

Follow-up

The day after treatment (day 1), a doctor will do a physical examination and we will do routine checks of your general well-being. We will also ask you about any other medications you take and any new symptoms or side effects you are experiencing. In addition, we will take blood for tests (approximately 4 teaspoons) and perform an ECG. You will still be on the ward for routine care, so we will perform the tests there. Taking part in the study will not mean you spend longer in hospital. If you are still in hospital the following day (day 2), we will repeat the same tests again.

After you have been sent home, we will ask you to come back to hospital on day 7 and day 30 after your treatment. We will pay your travel expenses for these follow-up visits, or if you prefer we can arrange for transport to take you to and from your visits. At each visit, you will have the same tests described above. If you are unable to come to the hospital and you agree to this, we might arrange for a nurse to visit you at home to perform the tests there. If you do not come to the hospital or see our nurse at home, we will contact you by phone to ask about any other medications you take and any new symptoms or side effects you are experiencing. We might also check your medical records and take note of any medications and medical events recorded there.

**Your involvement at a glance**

| **Visit name** | **When will it happen?** | **Where will I be?** | **What will happen?** |
| --- | --- | --- | --- |
| **Baseline/screening** | Day -1 or 0 | On the hospital ward for routine care | - routine medical checks (temperature, blood pressure, oxygen levels, breathing and pulse rate) - questions about your medical history and medications you take - ECG - blood sample |
| **Treatment** | Day 0 | Transfer to the Clinical Research Facility | - routine medical checks - questions about other medications you take, your symptoms and side effects - infusion of blood cells - regular safety checks - ECG - blood sample |
| **Follow-up 1** | Day 1 | On the hospital ward for routine care | - routine medical checks - questions about other medications you take, your symptoms and side effects - ECG - blood sample |
| **Follow-up 2 (optional)** | Day 2 | On the hospital ward for routine care (no visit if already discharged) |  |
| **Follow-up 3** | Day 7 | Travel to hospital for visit (or we can send a nurse to your home for the visit or we will contact you by phone) |  |
| **Follow-up 4** | Day 30 | Travel to hospital for visit (or we can send a nurse to your home for the visit or we will contact you by phone) |  |

**Is there anything I need to do or avoid?**

If you agree to take part in this study, you would not be able to join another clinical trial that tests a specific drug or cell treatment until the final follow-up for MAIL has been completed.

If you are a woman, you should not take part in this study if you are pregnant or are trying to get pregnant. You should also not breastfeed whilst taking part in the study. If you are a woman who can get pregnant, we will ask you to take a pregnancy test before you can take part.

If you decide to take part, you and your partner(s) must agree to use a highly effective method of contraception until 90 days after the end of the study. Highly effective birth control measures include hormonal contraception (combined or progestogen-only), intrauterine device or intrauterine hormone-releasing system (also known as coil), bilateral tubal occlusion (female sterilisation), vasectomy and sexual abstinence (no heterosexual intercourse during the entire period stated above; periodic abstinence, also known as natural family planning or the rhythm method, is not acceptable). If you or your partner become pregnant during the study you must notify the study team right away. For safety reasons, female participants who become pregnant will be discontinued from the study. We will ask for consent to check your (or your partner’s) medical records until 1-2 months after the due date of the baby to review the outcome of the pregnancy. These are normal precautions that are generally applied to all new treatments, because we do not yet know what effects, if any, the treatment may have on pregnancies.

**What are the possible benefits of taking part?**

We do not know if you will directly benefit from taking part in this trial. However, the information we get from this study will help improve the treatment of people who require treatment in the future.

**What are the possible disadvantages of taking part?**

This is a new treatment and we do not yet know for certain what adverse effects, if any, it may have. However, different types of macrophages have been given to humans with other conditions in studies before. These studies did not show any severe adverse effects from the macrophage treatment. The cells you receive are from a donor who has been screened to the same high standard as all blood donors.

You will be given an antihistamine (chlorphenamine or cetirizine) to reduce the likelihood of having an allergic reaction to the macrophage treatment. These antihistamines are routinely used to prevent and treat allergic reactions and the side effects are well known. The most common side effect of chlorphenamine is sedation varying from slight drowsiness to deep sleep, and the most common side effects of cetirizine are drowsiness and headaches.

You will be monitored closely during treatment and should report any side effects to your doctor or nurse. You will be given appropriate medication to treat any side effects that might occur. Rare, known side effects of cell therapies include the following symptoms:

- Nausea, vomiting, chest tightness, skin flushes and a rise in blood pressure and/or temperature. These side effects last only a short time and can be treated.
- Very rarely may cause a drop in blood pressure, a change in heart rate, difficulty in breathing or an acute allergic reaction. These side effects last only a short time and can be treated.

There is a chance (2-9 in 100) that having the cell treatment will cause you to produce antibodies against a protein called Human Leukocyte Antigen (HLA). HLA antibodies pose absolutely no risk to you. You will still be eligible to receive an organ transplant if required (such as a liver transplant in the unlikely setting of your liver function getting a lot worse or, for a reason unrelated to paracetamol, a kidney transplant at some time in the future). If you do develop HLA antibodies, it might be more difficult to find a suitable donor.

Attending the Day 7 and Day 30 follow-up appointments at the hospital will take up some of your time. The researchers involved in the study will do their best to work around you when organising appointments. We will pay your travel expenses for these follow-up visits, or if you prefer we can arrange for transport to take you to and from your visits.

You may experience some discomfort and bruising when having your blood samples taken.

**What if there are any problems?**

If you have a concern about any aspect of this study please contact Dr James Dear (0131 242 9214,) who will do his best to answer your questions.

If you were to become unable to make decisions for yourself during the trial (known as “loss of capacity”), then we would like you to continue taking part in the trial (assuming your nearest relative/legal representative is happy with this). We would continue to complete the follow-up visits and use your information (including blood samples) as normal.

In the unlikely event that something goes wrong and you are harmed during the research and this is due to someone’s negligence, then you may have grounds for a legal action for compensation against NHS Lothian but you may have to pay your legal costs. The normal National Health Service complaints mechanisms will still be available to you (if appropriate).

**What will happen if I don’t want to carry on with the study**

Your participation is entirely voluntary. You (or your legal representative if you were to lose mental capacity during the study) are free to withdraw from the study at any time and without giving a reason. Deciding not to take part or withdrawing from the study will not affect the healthcare that you receive, or your legal rights.

If you decide to withdraw, all information (including blood samples) you gave us before you left the trial will still be kept and used for the trial. If you withdraw after you received the cell product, we would like to continue collecting information about your health from your medical records. You can tell us if you do not want this to happen.

**What happens when the study is finished?**

After the study is finished, your data and blood samples will be stored for 30 years as required by law for this type of study. After this period, they will be disposed of securely.

We might make the study data available for other researchers to look at (in the UK and abroad). Before we make it available, we will make sure it does not contain any of your personal data so that you will not be identifiable.

With your permission, your anonymised blood samples will be stored for future research projects. When we test blood samples in the future we may send them to other academic institutions or commercial companies (in the UK or abroad) to help us run those tests. Your blood will only be used in future studies that have relevant permissions in place.

**Will my taking part be kept confidential?**

As routine, we will inform your GP about your hospital admission and that you took part in this study. All the information we collect during the course of the research will be kept confidential and there are strict laws which safeguard your privacy at every stage.

**How will we use information about you?**

We will need to use information from you and from your medical records for this research project.  This information will include your name, CHI number (a unique number that identifies you and is used by NHS Scotland for health care purposes), date of birth, ethnicity, gender and phone number.  People will use this information to do the research or to check your records to make sure that the research is being done properly. We will keep all information about you safe and secure (paper files and blood samples in locked facilities and electronic data on a secure database with restricted access).

People who do not need to know who you are will not be able to see your name or contact details. Your data will have a code number instead.

Once we have finished the study, we will keep some of the data so we can check the results. We will write our reports in a way that no one can work out that you took part in the study.

We will pass some of your personal information (including your name, CHI number and date of birth) on to the Scottish National Blood Transfusion Service. They require this information under the terms of the UK Blood Safety and Quality Regulations and will store it securely. Some of your personal information (age in years, ethnicity, gender and sex assigned at birth) will be entered onto the secure trial database, which is managed by the University of Edinburgh Clinical Trials Unit, to do the research.

### What are your choices about how your information is used?

- You can stop being part of the study at any time, without giving a reason, but we will keep information about you that we already have.
- If you choose to stop taking part in the study after you received the study treatment, we would like to continue collecting information about your health from your medical records. If you do not want this to happen, tell us and we will stop.
- We need to manage your records in specific ways for the research to be reliable. This means that we won’t be able to let you see or change the data we hold about you.
- If you agree to take part in this study, you will have the option to take part in future research using your blood samples saved from this study.

### Where can you find out more about how your information is used?

You can find out more about how we use your information

- at [www.hra.nhs.uk/information-about-patients/](https://www.hra.nhs.uk/information-about-patients/)
- our leaflet available from [[**www.hra.nhs.uk/patientdataandresearch**](http://www.hra.nhs.uk/patientdataandresearch)
- by asking one of the research team (Dr James Dear, 0131 242 9214,), or

**What will happen to the results of the study?**

This study will be written up and submitted for publication in a medical journal. It is likely that the results will also be presented at academic meetings or conferences. Once the study has been published a summary of the findings will be available on the Edinburgh Clinical Trials website (https://www.ed.ac.uk/usher/edinburgh-clinical-trials/our-studies/all-current-studies/macrophage-therapy-for-acute-liver-injury-mail-tri). You will not be identifiable from any published results.

**Who is organising and funding the research?**

This study is led by Professor James Dear, Personal Chair of Clinical Pharmacology at the University of Edinburgh, and managed by the Edinburgh Clinical Trials Unit.

The study is being funded by the Medical Research Council (MRC) and sponsored by University of Edinburgh and NHS Lothian. Some of the research team’s salaries are funded by the MRC award for this study.

**Who has reviewed the study?**

The study proposal has been reviewed by experienced researchers at the MRC, the University of Edinburgh, the Scottish National Blood Transfusion Service and in the NHS. All research in the NHS is looked at by an independent group of people called a Research Ethics Committee. A favourable ethical opinion has been obtained from the North East – York Research Ethics Committee. The study has also been approved by the Medicines & Healthcare products Regulatory Agency (MHRA). NHS Management Approval has also been given.

**Researcher Contact Details**

If you have any further questions about the study please contact:

Dr James Dear

Out of hours helpline:

**Independent Contact Details**

If you would like to discuss this study with someone independent of the study please contact Professor Michael Eddleston (0131 242 1383,).

**Complaints**

If you wish to make a complaint about the study please contact:

Patient Experience Team

2 – 4 Waterloo Place, Edinburgh, EH1 3EG

0131 536 3370

**CONSENT FORM**

**Macrophage Therapy for Acute Liver Injury (MAIL) Trial**

|  | Please **initial** box |
| --- | --- |
| 1. I confirm that I have read and understand the information sheet (V3.0 11May2023) for the above study. I have had the opportunity to consider the information, ask questions and have had these questions answered satisfactorily. |  |
| 2. I understand that my participation is voluntary and that I am free to withdraw at any time, without giving any reason and without my medical care and/or legal rights being affected. |  |
| 3. I give permission for the research team to access my medical records for the purposes of this research study. |  |
| 4. I understand that relevant sections of my medical notes and data collected during the study may be looked at by individuals from the Sponsor (University of Edinburgh and/or NHS Lothian), from regulatory authorities or from the NHS organisation where it is relevant to my taking part in this research. I give permission for these individuals to have access to my data and/or medical records. |  |
| 5. I give permission for my personal information (including my name, CHI number, date of birth, ethnicity, gender, sex assigned at birth and telephone number) to be collected and stored in NHS Lothian. |  |
| 6. I give permission for some of my indirectly identifiable personal information (age in years, ethnicity, gender and sex assigned at birth) to be entered onto the secure trial database, which is managed by the University of Edinburgh. |  |
| 7. I give permission for some of my directly identifiable personal information to be passed to the Scottish National Blood Transfusion Service (name, CHI number and date of birth). |  |
| 8. I agree to my General Practitioner being informed of my participation in the study. |  |
| 9. I give permission for any previous blood samples stored at the Royal Infirmary of Edinburgh to be obtained from the laboratory and analysed for this study. |  |
| 10. I agree to my anonymised data and blood samples being used in future approved studies (academic and commercial, UK and abroad): | Yes No |
| 11. I agree to take part in the above study. |  |

| Name of Person Giving Consent |  | Date |  | Signature |
| --- | --- | --- | --- | --- |
| Name of Person Receiving Consent |  | Date |  | Signature |

1x original – into Site File; 1x copy – to Participant; 1x copy – into medical record
